# A cross-sectional study of low birth satisfaction among Iranian postpartum women during COVID-19 epidemics’ fifth wave

**DOI:** 10.1101/2022.09.08.22279714

**Authors:** Forough Mortazavi, Maryam Mehrabadi

## Abstract

**Background:** Birth dissatisfaction may increase the risk for postpartum depression and requests for an elective cesarean for the next birth. The outbreak of COVID-19 pandemic has had a considerable impact on the healthcare systems and their users in many aspects. We investigated predictors of birth satisfaction in a sample of Iranian postpartum women during the COVID-19 epidemics’ fifth wave.

**Methods:** This cross-sectional study was conducted on 601 postpartum women admitted to postpartum wards of Mobini maternity hospital using a convenience sampling method between 2 Aug and 18 September 2021. We collected data on socio-demographic, obstetric, labor and birth, and psychological variables. We used the general linear model and multiple linear regression analyses to determine predictors of birth satisfaction.

**Results:** The mean birth satisfaction score was 28.6±7.3. The percentages of mothers who gave birth by elective and emergency cesarean were 19.5% and 10.8%, respectively. Overall predictors of birth satisfaction were emergency cesarean [-7.463(-9.310, -5.616), instrumental birth [-3.571(-6.907, -0.235)], episiotomy [-2.227 (-3.591, -0.862)], Entonox analgesia [-1.548(-2.726, -0.371)], Well-being score < 50 [-1.812(-3.146, -0.478)], fear of COVID-19 [-1.216(-2.288,, -0.144)], low satisfaction with pregnancy -2.539(-3.952, -1.127) and low satisfaction with spouse’s support [-2.419(-4.598, -0.240)].

**Conclusions:** During the pandemic, fear of COVID-19, low level of well-being, low satisfaction with pregnancy and low satisfaction with spouse’s support as well as women’s experience of emergency cesarean, instrumental birth, episiotomy, and Entonox analgesia, are exerting negative influences on birth satisfaction. To improve birth satisfaction and thus maternal mental health interventions to lower fear of contracting COVID-19 and reduce rates of episiotomy, emergency cesarean, and instrumental birth are recommended.

## Introduction

Childbirth is a turning point in a woman’s life that can be either a wonderful experience or a traumatic one. Although the primary goal of childbirth is the birth of a healthy infant from a healthy mother, women would generally like it to be a positive and satisfying experience (1). Birth satisfaction refers to the mother’s overall assessment of childbirth and the extent to which she perceived it as a positive and satisfying experience (2). Studies in developed countries report that 7–10% of women have had a negative birth experience (3).

Several studies have found positive association between birth dissatisfaction and a number of outcomes including postpartum depression (2-4), difficulties in mother-baby bonding, poor maternal care of the baby and abstaining from exclusive breastfeeding (3). Findings of a study in New York City hospitals indicate that women with lower birth satisfaction were more likely to experience higher levels of anxiety, stress, and depressive symptoms during the postpartum period (4). Birth dissatisfaction may also increase requests for an elective cesarean for the next birth (5) and the likelihood of mothers delaying their next pregnancy (6). Dissatisfaction with vaginal birth among women in Hong Kong after their first childbirth resulted in 23.8% of them changing their preferred mode of birth from vaginal to elective cesarean (5).

Several factors have been found to contribute to dissatisfaction with childbirth including emergency cesarean (5, 7, 8), elective cesarean, higher family income, use of epidural analgesia (5), induction, unwanted pregnancy (7), primiparity, low level of well-being, low satisfaction with pregnancy, severe fear of childbirth, long time between hospital admission and giving birth (8), and lack of support from partner (7, 8). Also, childbirth satisfaction has been found to be associated with planned childbirth (9), mothers’ acquaintance with labor, moderate labor pain (10), women’s participation in deciding matters that affect them (11), and empathy and respectful behavior of caregivers (11, 12).

The COVID-19 pandemic has led to increasing rates of anxiety, depression, and stress among women during pregnancy and childbirth (13-15). Stress was related to constrained antenatal care during the pandemic, worries about being infected with the virus, the possibility of negative effects of the infection on the baby, and worry about the health of loved ones (16). Fear of COVID-19 has been found to be a common problem during the pandemic with a significant impact on women’s well-being (13). It had a statistically significant relationship with depression, suicidal tendencies, and poor psychological quality of life in pregnant women (17). Studies have also found that, just as in the pre-pandemic period, dissatisfaction with childbirth during COVID-19 pandemic is associated with postpartum depression (15, 18). The pandemic has also impacted the organization and the practices of health care provision in the antenatal, intrapartum, and postnatal periods. It has also impacted the attitudes and the responses of healthcare users. Factors that might have been impacted include pregnancy intention, care providers’ behavior towards women, more restrictive hospital rules, the likelihood of experiencing a hassle-free pregnancy, and request for elective cesarean. It may also have increased households’ economic concerns. Therefore, it would not be unreasonable to expect lower levels of birth satisfaction in women giving birth during COVID-19 pandemic (15).

Like many other countries, Iran has experienced high rates of cesarean (19) and maternal requests for cesarean (20) in recent decades. At the same time, the decreasing total fertility rate (TFR) has been increasingly viewed as a threat to the future development of the country and thus a cause for concern (21). Because birth dissatisfaction may increase requests for an elective cesarean for the next birth and the likelihood of mothers delaying their next pregnancy, there is concerns that the COVID-19 epidemic with its negative impact on birth satisfaction may undermine the Health Transformation Plan (HTP), an initiative launched by the Iranian government for the reform of the healthcare system in 2014 (22). HTP includes a number of guidelines and measures with regard to maternity care aiming to promote maternal experience of childbirth. Thus, we hypothesize that maternal birth experience and childbirth satisfaction might have been affected by COVID-19 pandemic. In the present study, we investigated the extent to which fear of COVID -19, maternal well-being status, and labor and birth factors predicted lower birth satisfaction during the COVID-19 epidemics’ fifth wave in Sabzevar, Northeast Iran.

## Materials and Methods

### Design, participants, and data collection

This cross-sectional study was conducted from Aug 2 to September 18, 2021. Women were recruited in the postpartum and surgery wards of Mobini Hospital affiliated with Sabzevar University of Medical Sciences. As a matter of hospital policy, they were usually hospitalized for the first 24 hours and 48 hours after vaginal delivery and cesarean, respectively. Women who had vaginal deliveries completed the 45-item study questionnaire on the morning of the day after childbirth when they felt they were ready for the task. In the cesarean group, they filled out the scales on the morning of the second day after birth. Sampling was done using the convenience method. Women who gave birth to a healthy, live infant were included in the study. We excluded women with mental illness under treatment, women with severe postpartum complications, women with infants admitted to the intensive care unit, those with preterm birth, and COVID-positive women. Women admitted for birth in the hospital who showed mild signs of COVID were tested and those with positive results were isolated from other women in labor and postpartum. Women with severe disease were transferred to another facility dedicated to COVID-19 patients. The average annual birth rate in the hospital which was 5898 before the COVID -19 outbreak, decreased to 5245 in the first year after the outbreak. There were 4020 births in the first 9 months of the second year of the pandemic. A research colleague, who was a midwifery graduate, identified eligible mothers and, if they agreed to participate in the study, presented them with written consent forms and anonymous questionnaires. We trained the midwife on how to present the questionnaire and collect the data. Demographic, social, and obstetric information of the mothers were extracted from their medical records and recorded by the colleague.

### Ethical consideration

The Ethics Committee of Sabzevar University of Medical Sciences has reviewed and approved this study (approval number: IR.MEDSAB.REC.1399.103). All procedures were performed in accordance with the guidelines of Sabzevar University of Medical Sciences, which is in accordance with the Declaration of Helsinki. Women who consented to participate in the study signed an informed consent form and were assured about the confidentiality of their information. After recording the socio-demographic information of the women in the questionnaire, the midwife asked women to fill out the scales. The questionnaires and scales were anonymous.

### Instruments

#### Interview form

A three-part questionnaire was completed by the midwife. The first part contained questions on socio-demographic characteristics (including age, education, residency, and job). The second part consisted of obstetrical information (such as parity, attending prenatal classes, history of chronic disease, the desirability of pregnancy, poor obstetric history, infant gender, history of abortion, and complicated pregnancy). The third part consisted of labor and birth information (such as mode of birth, induced or spontaneous labor pain, pain relief method during labor and birth, having a private midwife at birth, gestational age, birth weight, admission to birth duration, and episiotomy/tear repair).

The level of satisfaction with pregnancy was assessed based on the extent of health problems experienced during pregnancy using a five-point Likert scale from 1 to 5 (1 = dissatisfied to 5 = very satisfied). Women’s satisfaction with husband’s emotional/financial support and marital/sexual satisfaction were evaluated with the same scale. A question on receiving fundal pressure was also included. Women rated their household income as 1= insufficient or 2= sufficient. (Recorded in a supplementary file 1)

To measure birth satisfaction, there are two scales developed for measuring birth experience: the Birth Satisfaction Scale–Revised (BSS-R) and the Childbirth Experience Questionnaire (CEQ2) (23). We examined both scales and found similar results with regard to their ability to explain birth satisfaction variance. We present our results on the CEQ2 in a supplementary file.

#### The Birth Satisfaction Scale–Revised (BSS-R)

The Birth Satisfaction Scale–Revised (BSS-R), created by Martin & Martin in 2014, is regarded as the best instrument for measuring women’s birth satisfaction (24). The BSS-R includes 10-items in three factors: stress experienced during labor, women’s personal attributes, and quality of care provision. Participants were asked to rate each item on a four-point Likert scale which ranges from 0-4 (0 = strongly disagree to 4 = strongly agree). The BSS-R showed acceptable internal consistency (Cronbach’s alpha = 0.79). Its total score ranges from zero to 40. Mortazavi and colleagues translated the scale into Persian. The validity study of the Persian BSS-R indicted that it has three dimensions which are identical to those proposed by the developers of the original scale. The reliability of the Persian BSS-R has been confirmed (Cronbach’s alpha = 0.76) (25). In the present study, we calculated a Cronbach’s alpha value of 0.734.

#### The World Health Organization’s Well-Being Index (WHO-5 Well-Being Index)

The 5-item World Health Organization Well-Being Index (WHO-5) assesses the emotional well-being of individuals over the preceding two weeks. It has five items which are rated using a 6-point Likert scale where zero represents ‘having good feelings at no time’ and five represent ‘having good feelings all the time’ (26). The scale’s total score ranges from 0 to 25 which is converted to a scale of 0 to 100. The scale is used in screening programs for depression with a score of 50 as the cut-off point. Individuals with scores less than fifty should be referred for further assessment. The validity and reliability of the Persian version of WHO-5 in pregnant women were confirmed and its unidimensionality and reliability was proven (Cronbach’s alpha = 0.85) (27). In our study, we calculated a Cronbach’s alpha value of 0.889.

#### Fear of COVID-19 Scale (FCV-19S)

The Fear of COVID-19 Scale (FCV-19S) is one of the most widely used scales for assessing fear of COVID-19. It was developed by Ahorsu et al. (2020) for the purpose of assessing fear of COVID-19. The scale was translated into Persian and the validity study for the scale was conducted using a sample of Iranian students (28). It is a unidimensional instrument consisting of 7 items rated on a 5-point Likert-type scale (1 to 5 points). The total score ranges from 7 to 35 with higher scores indicating a higher level of fear of COVID-19. A Cronbach’s alpha value of 0.88 was reported in the original study on the scale (28). In our study, we calculated a Cronbach’s alpha value of 0.928.

### Data analysis

We used the SPSS version 18 to analyze the data. Descriptive statistics were used to characterize the participants. We evaluated the normal distribution of birth satisfaction scores and other quantitative variables using skewness and kurtosis. We used the general univariate linear model to identify independent variables with a significant impact on birth satisfaction scores. Then, variables with p <0.25 in the univariate linear regression were entered into five separate multiple linear regression analyses by the backward-LR method. These analyses enabled us to determine the demographic, obstetric, labor and birth, psychological, and overall predictors of birth dissatisfaction. We checked linear regression assumptions. The normality of residuals was verified and collinearity statistics indicated no multicollinearity (tolerance < 1 and variance inflation factor < 2). The effect sizes for the entire model and main predictors of birth satisfaction scores were calculated.

## Results

Of the 676 women who gave birth in Mobini hospital during the study period, 67 women were excluded from the study of whom two women were COVID positive, 27 had a preterm birth, 35 women had an infant in NICU, four women experienced severe postpartum hemorrhage, and eight women did not consent to participate in the study. Overall, 601 women participated in the study. The mean value of age (year), education (years), gestational age (week), birth weight (gr), and admission to delivery duration (hour) were 28.7±6.6, 11.1±4.1, 39.1±1.2, 3250±465, and 7.2 ±8.0, respectively. The mean scores of FVC-19S, WHO-5, and birth satisfaction were 14.7±7.5, 69.5±26.8, 67.5±13.0, and 28.6±7.3, respectively. Sixty-five point nine percent (65.9%) of the participants were multiparous. The correlation between birth satisfaction total scores and the duration between admission to hospital and giving birth was 0.247 (p<0.001). Sample characteristics and the results of general linear models for the relationship between the BSS-R scores and independent variables are presented in table1.

**Table 1.**
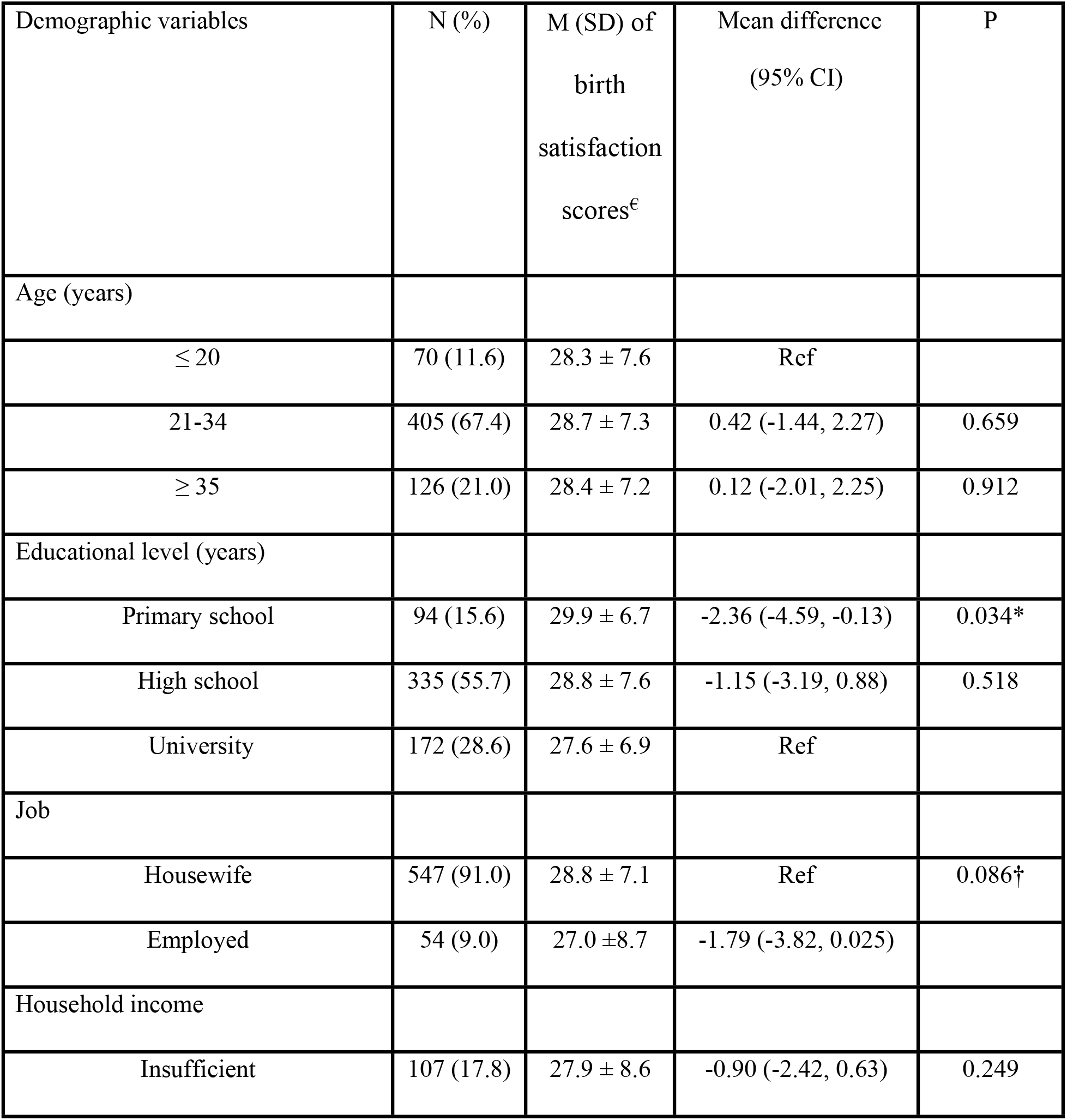
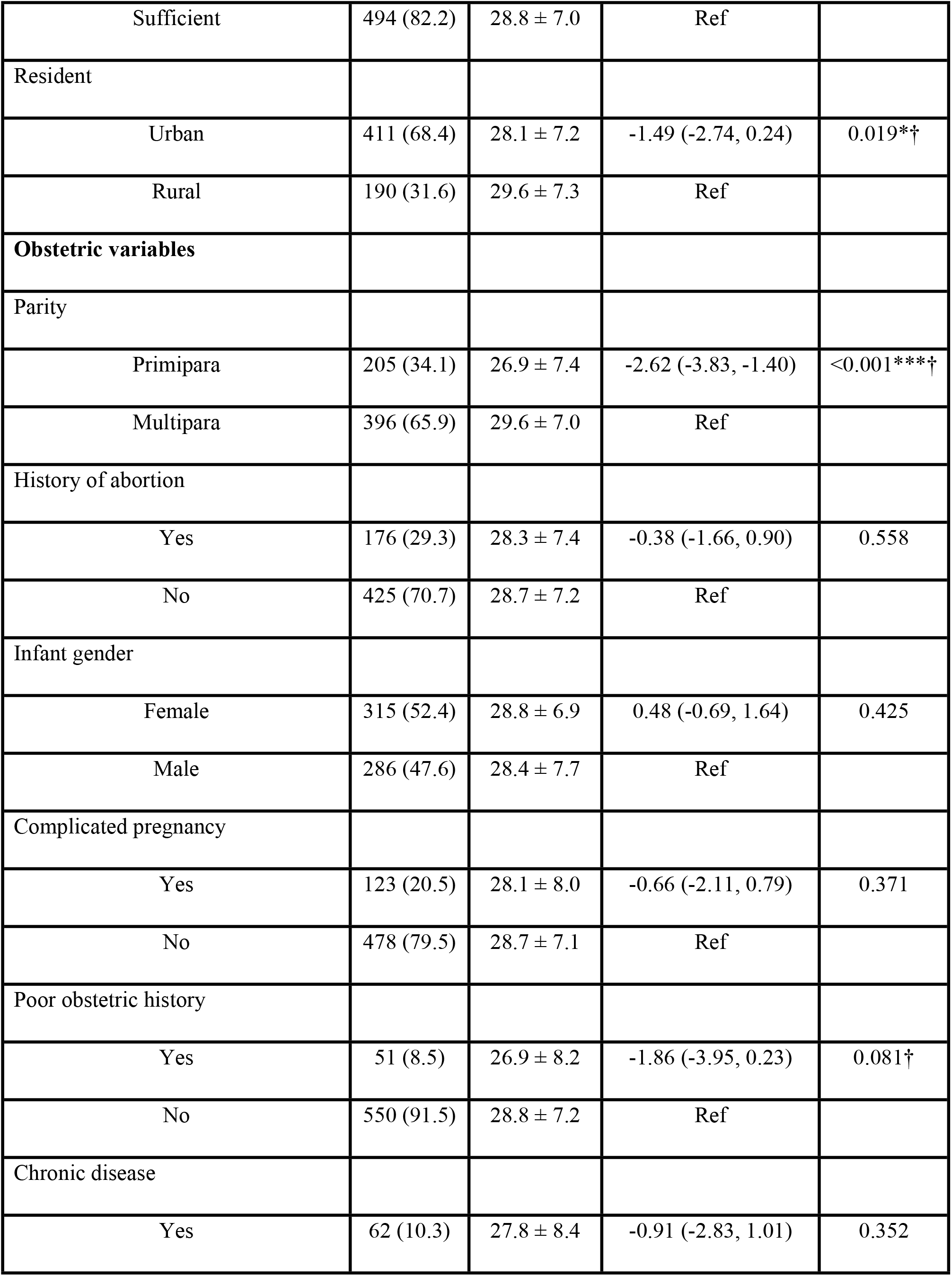
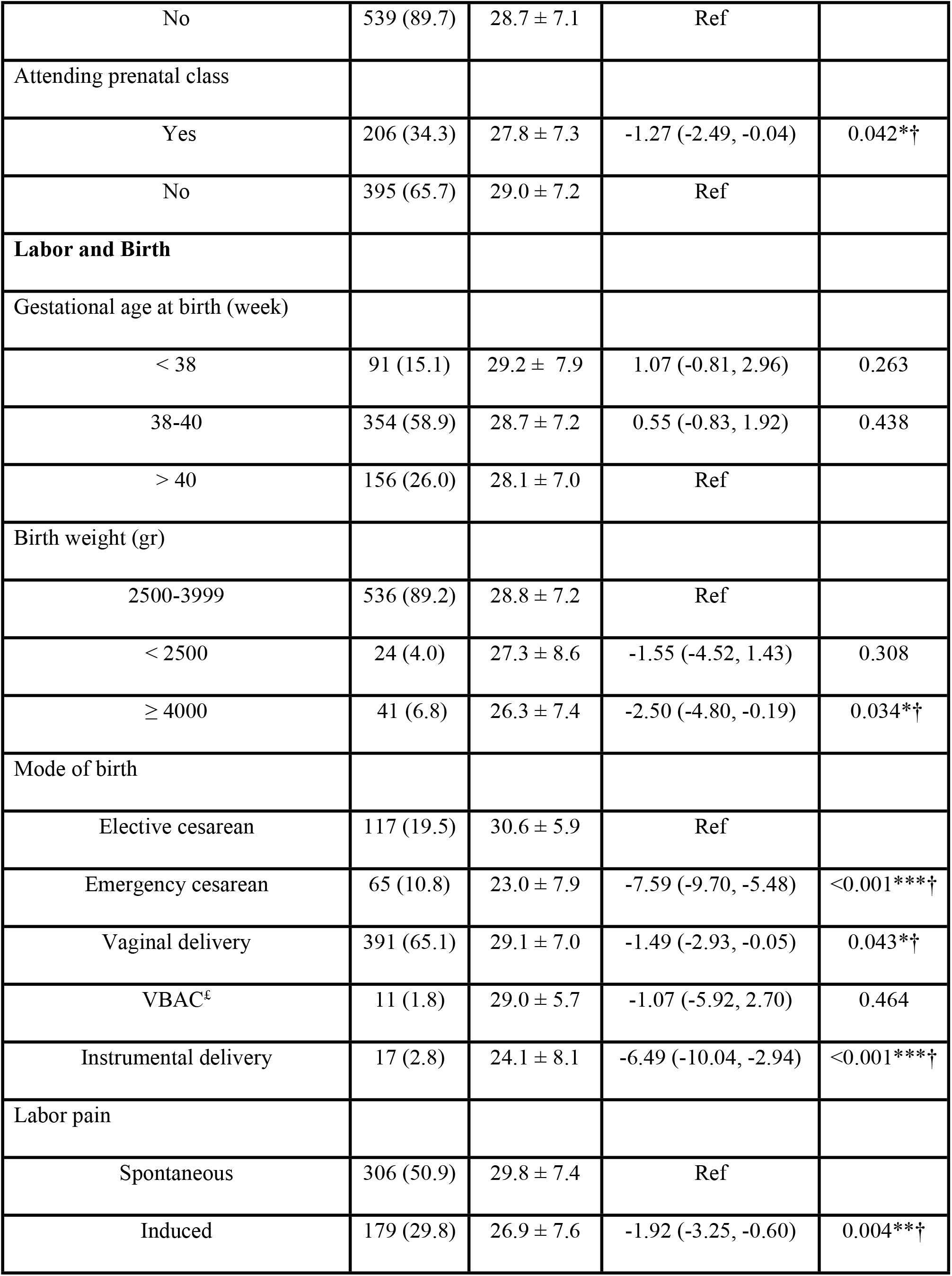
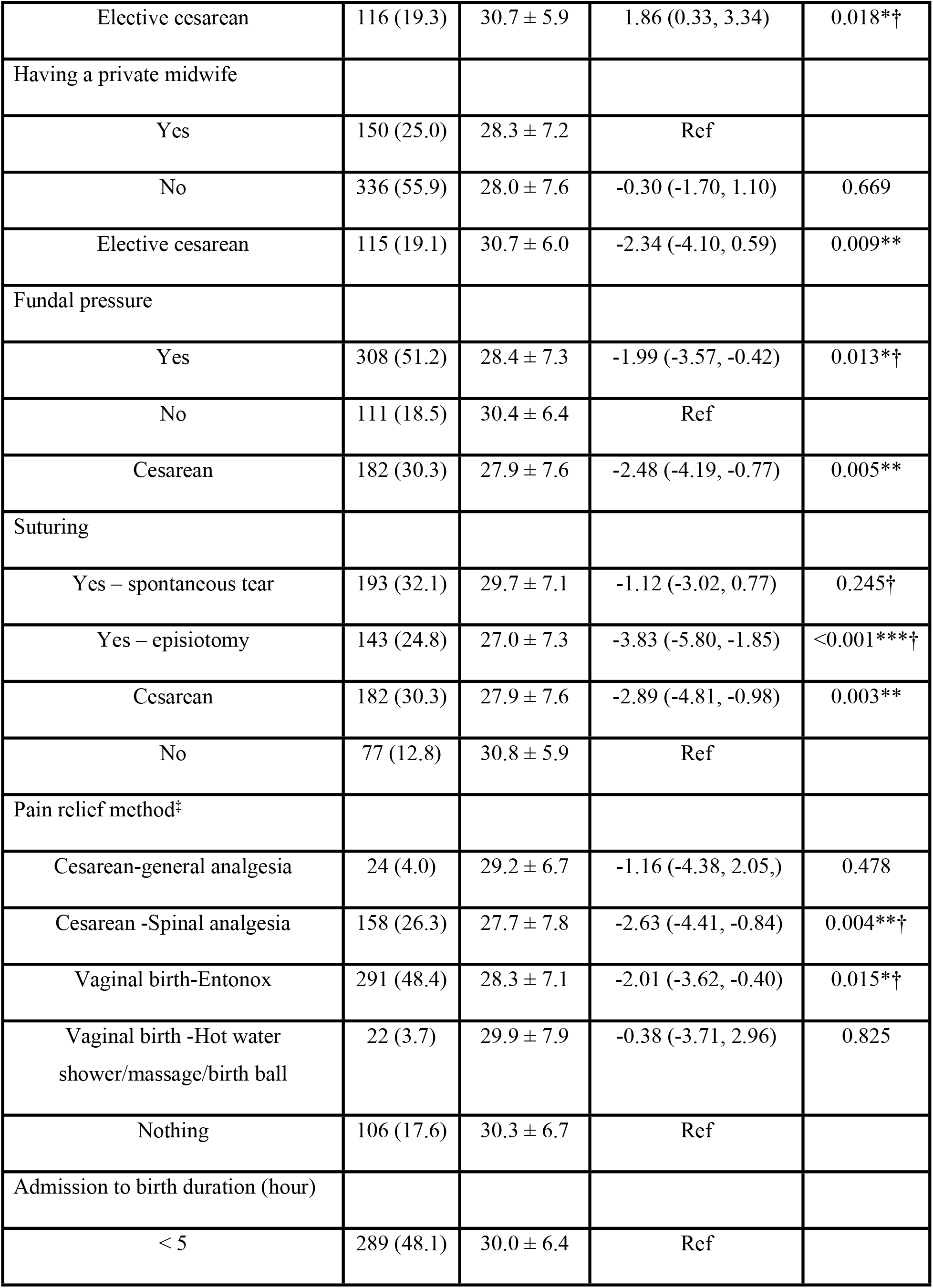
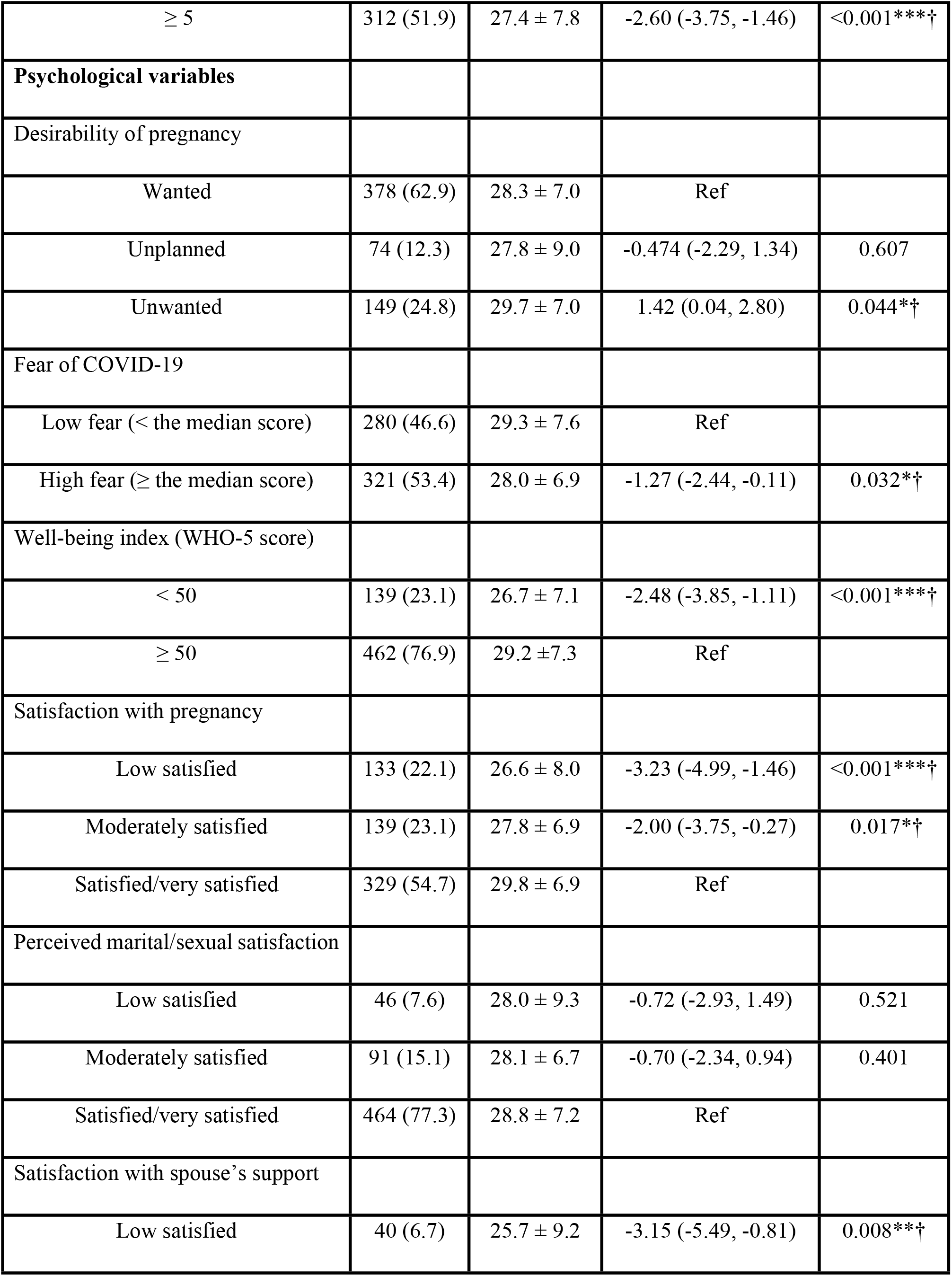
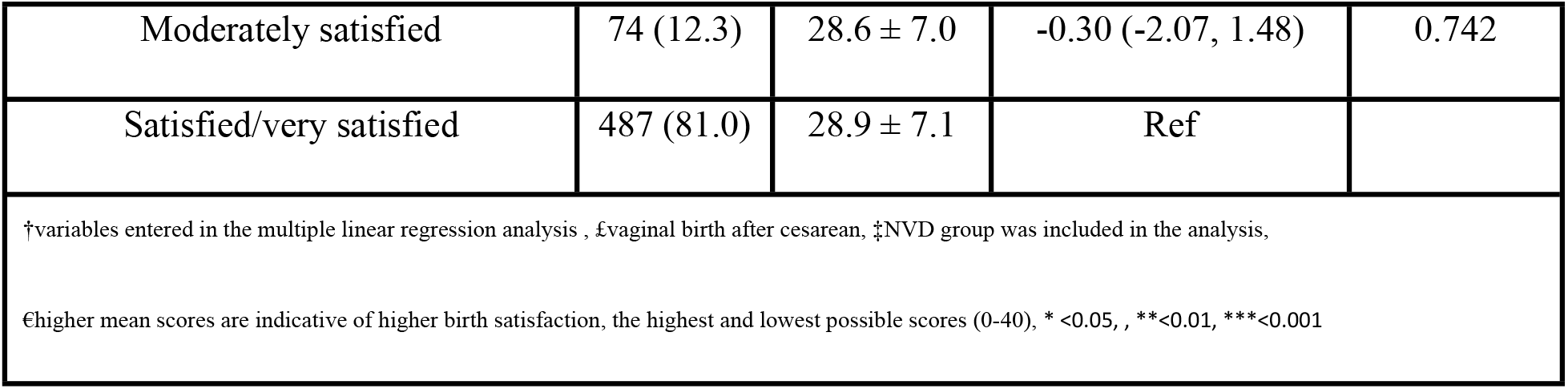
Sample characteristics, mean (SD) of birth satisfaction scores, and the results of general linear models on birth satisfaction scores (N = 601).

In table 2, the results of multiple linear regression analysis for the birth satisfaction scores are presented. Also, effect sizes for the main predictors of birth satisfaction are included in table 2. Obstetric and labor and birth predictors of birth satisfaction were primiparity [B = -2.650, CI (-3.861, -1.439)], birth weight (gr) ≥ 4000 [-2.351(-4.521, -0.180)], emergency cesarean [-7.332 (-9.276, -5.389)], and episiotomy [-2.466 (-3.853, -1.079)]. Psychological predictors of birth satisfaction were unwanted pregnancy [2.292 (.958, 3.625)], Well-being score < 50 [-1.742 (-3.145, - 0.338)], low satisfaction with spouse’s support [-2.523 (-4.828, -0.219)], and low [-2.910 (-4.400, -1.419)] and moderate [-2.168 (-3.580, -0.755)] satisfaction with pregnancy. Overall predictors of birth satisfaction were emergency cesarean [-7.463(-9.310, -5.616), instrumental delivery [-3.571(-6.907, -0.235)], episiotomy [-2.227-3.591, -0.862)], Entonox analgesia [-1.548(-2.726, -0.371)], Well-being score < 50 [-1.812(-3.146, -0.478)], fear of COVID -19 [-1.216(-2.288,, -0.144)], low satisfaction with pregnancy -2.539(-3.952, -1.127) and low satisfaction with spouse’s support [-2.419(-4.598, - 0.240)]. The overall proportion of the variance in birth satisfaction explained by all variables is 17.4% (effect size = 0.174). Labor and birth variables explained 12.2% of the variance in birth satisfaction.

**Table 2.**
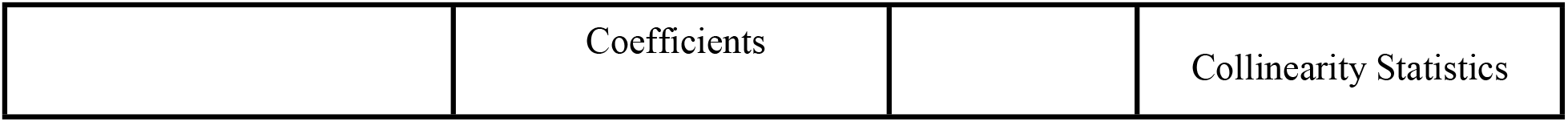
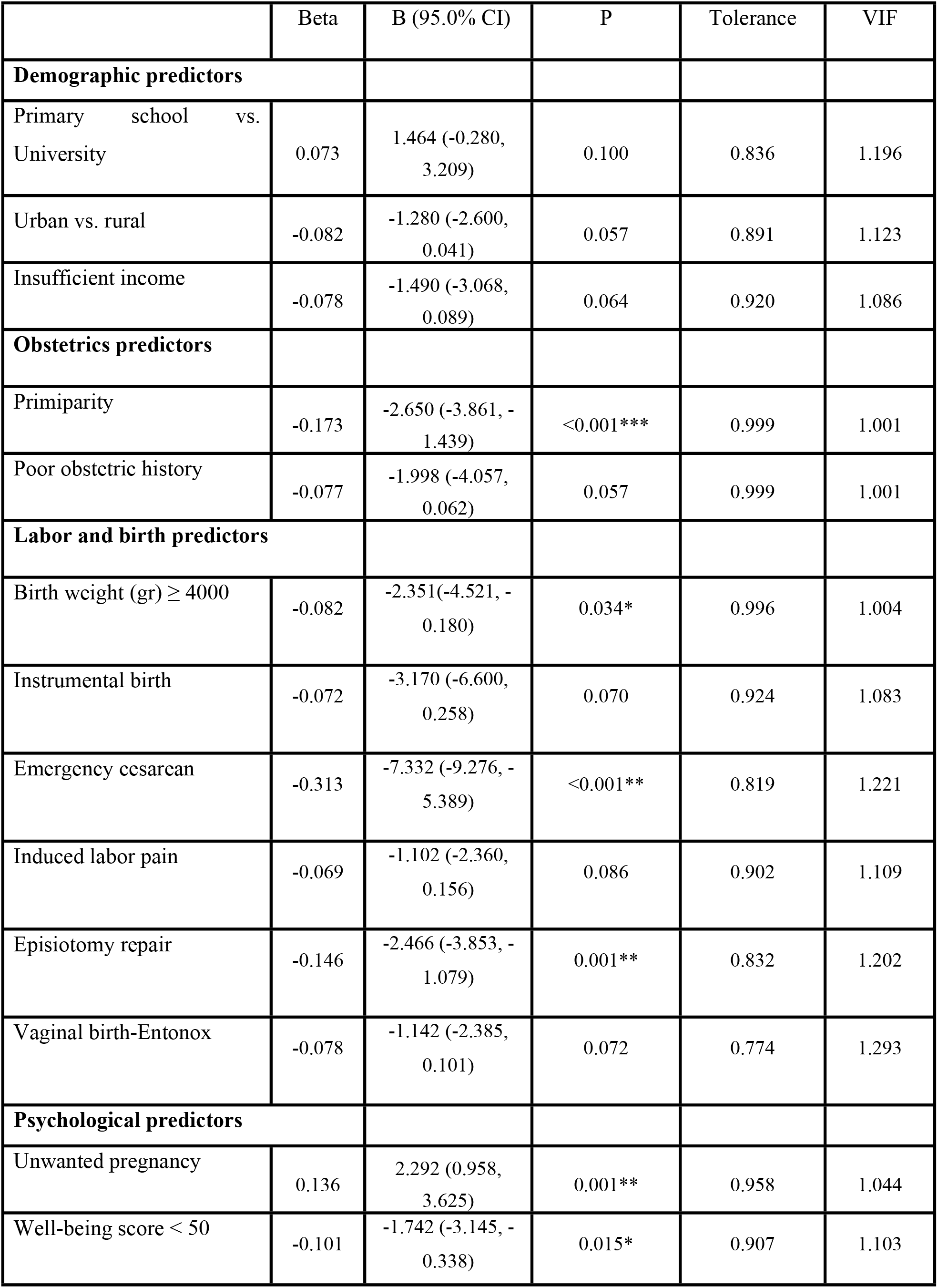
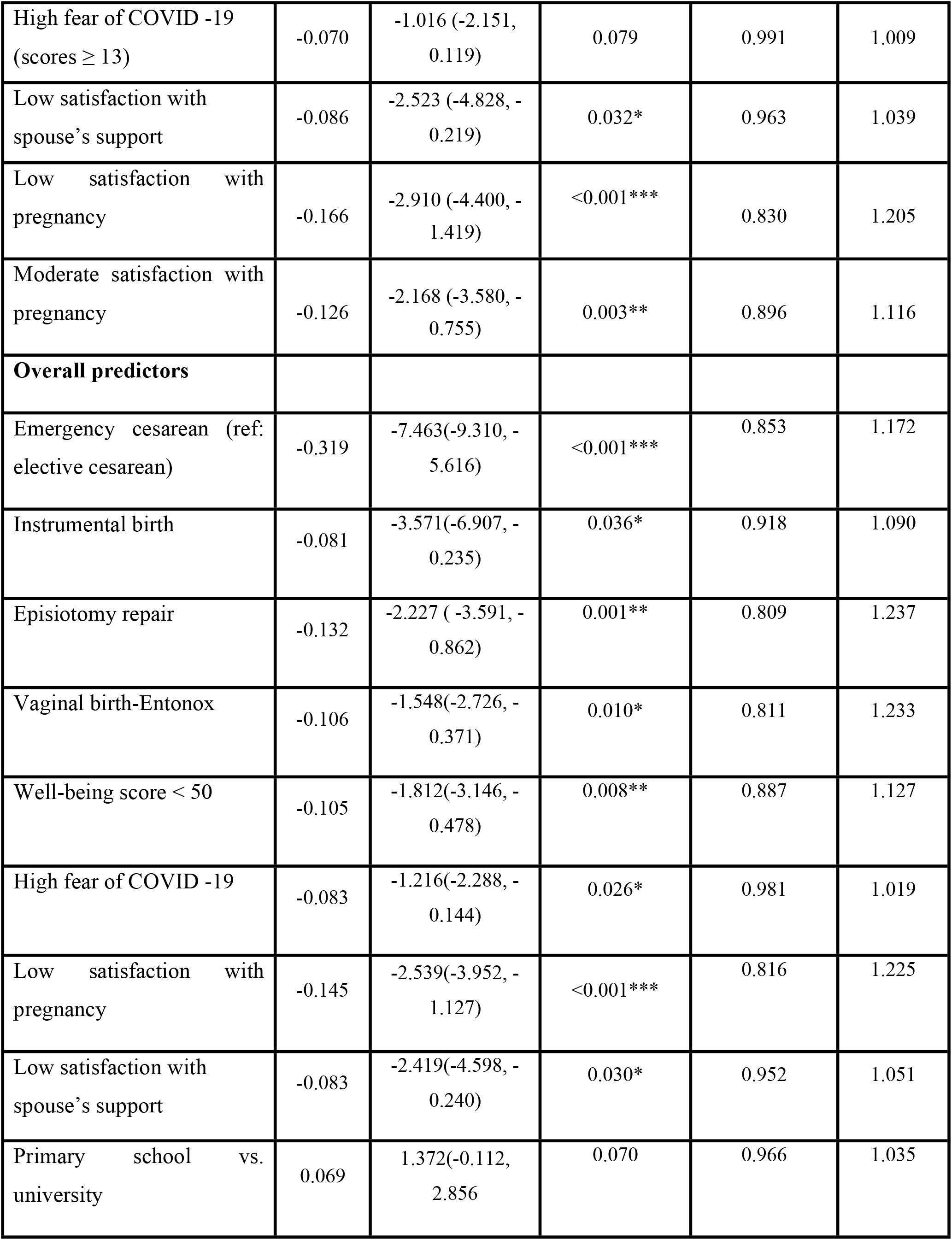
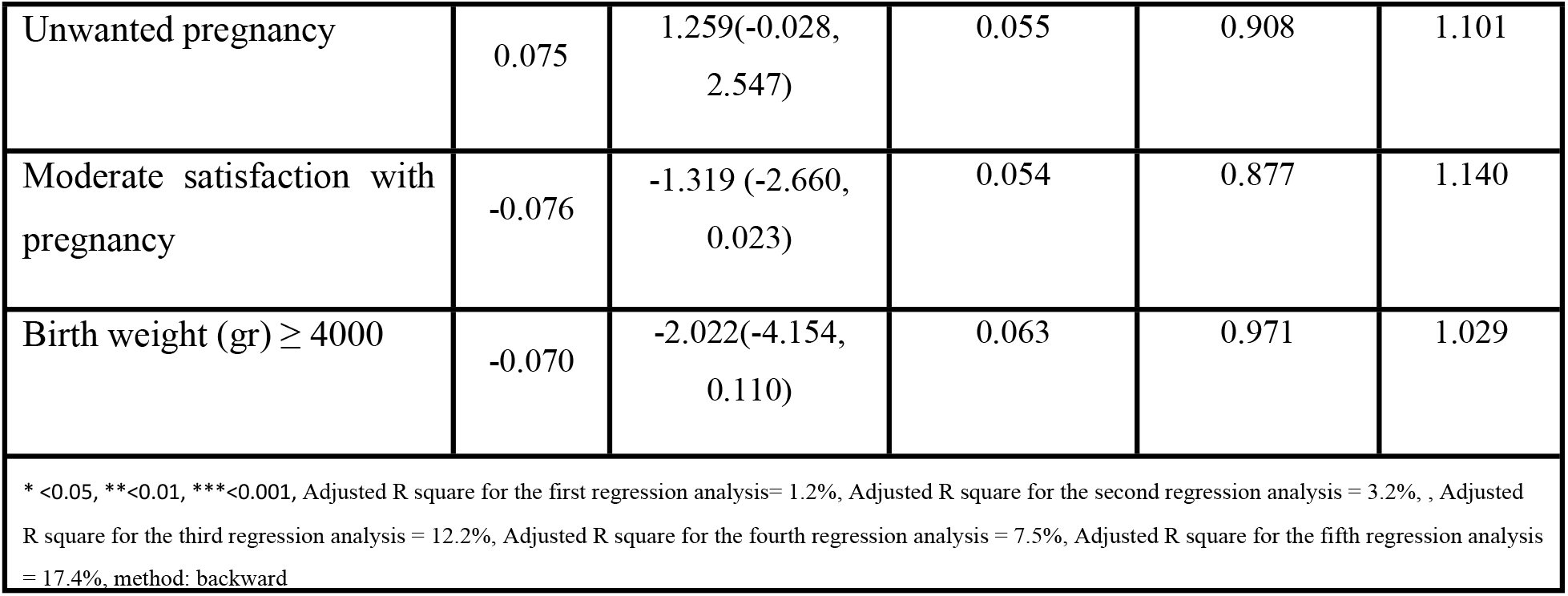
Results of multiple linear regression analysis on the birth satisfaction scores.

We investigated the relationship between the presence of a private midwife during labor and fear of COVID-19 in the case of women who had a vaginal delivery. Women who were accompanied by a private midwife reported a higher level of fear of COVID-19 than those without a private midwife (p = 0.044). The presence of a private midwife during labor had no relationship with birth satisfaction scores.

In table 3, the distribution of psychological variables including birth satisfaction, well-being, and fear of COVID-19 according to parity and household income in the pandemic period are presented. The BSS-R mean scores was higher in multiparas than primiparas (p<0.001). The FVC-19S and WHO-5 mean scores were not different between multiparas and primiparas (p > 0.05). They were also not different between women with sufficient and insufficient income (p > 0.05).

**Table 3.**
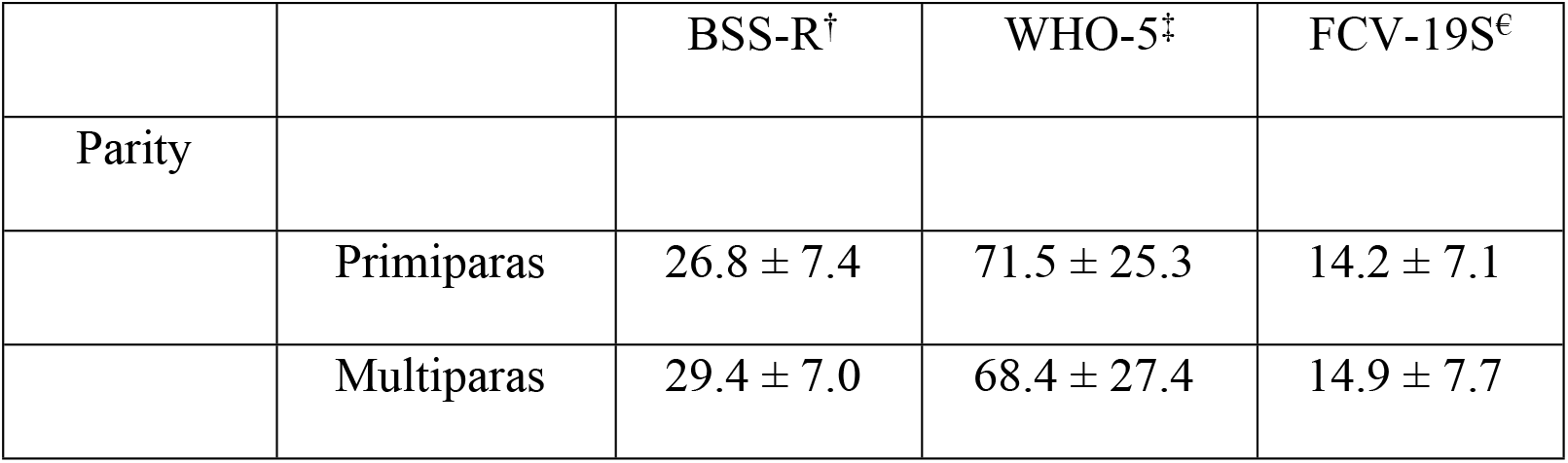
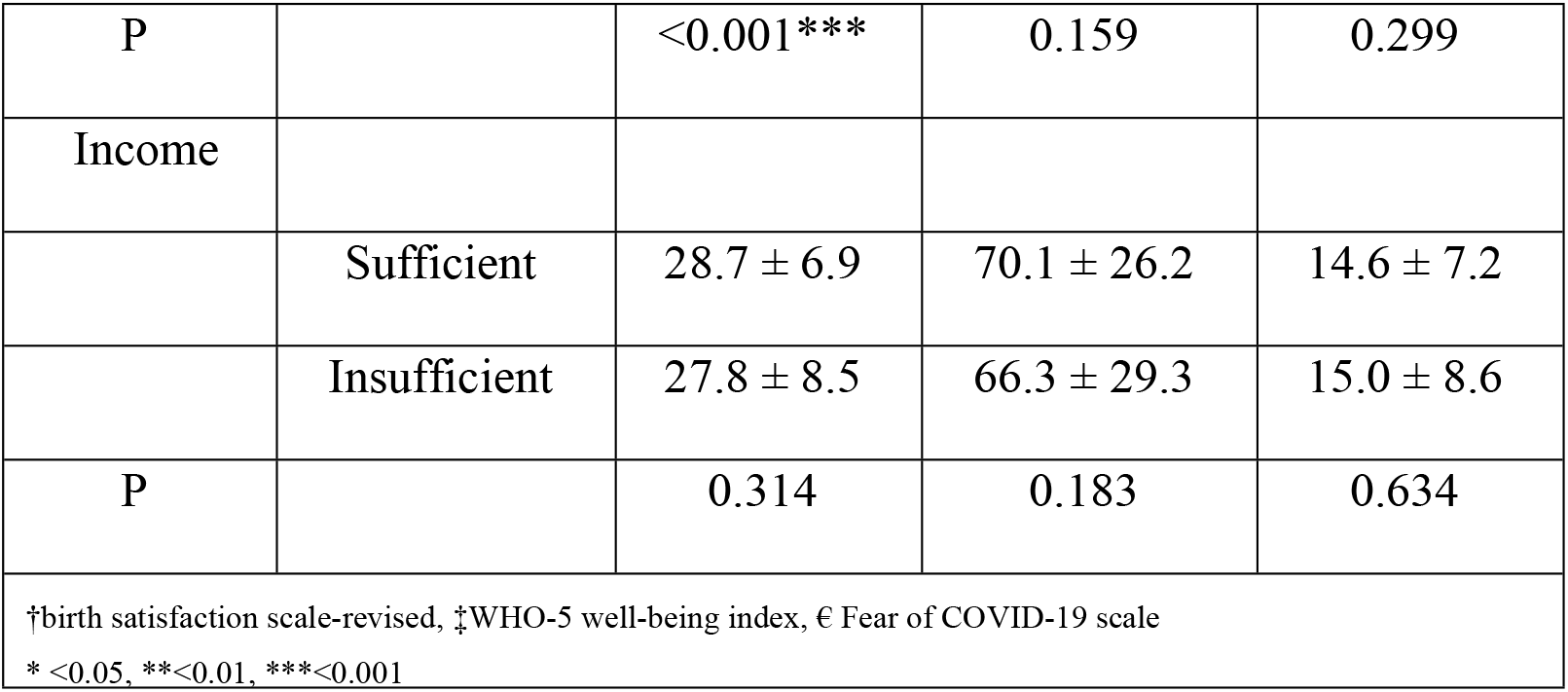
Mean scores of scales used in the study for different income and parity groups

|  |  | BSS-R<sup>†</sup> | WHO-5<sup>‡</sup> | FCV-19S<sup>€</sup> |
| --- | --- | --- | --- | --- |
| Parity |  |  |  |  |
| | Primiparas | 26.8 ± 7.4 | 71.5 ± 25.3 | 14.2 ± 7.1 |
| | Multiparas | 29.4 ± 7.0 | 68.4 ± 27.4 | 14.9 ± 7.7 |
| P |  | <0.001*** | 0.159 | 0.299 |
| Income |  |  |  |  |
|  | Sufficient | 28.7 ± 6.9 | 70.1 ± 26.2 | 14.6 ± 7.2 |
|  | Insufficient | 27.8 ± 8.5 | 66.3 ± 29.3 | 15.0 ± 8.6 |
| P |  | 0.314 | 0.183 | 0.634 |
| †birth satisfaction scale-revised, ‡WHO-5 well-being index, € Fear of COVID-19 scale<br>* <0.05, **<0.01, ***<0.001 |  |  |  |  |

## Discussion

We investigated the predictors of birth satisfaction in early postpartum during the COVID-19 epidemics’ fifth wave in Iran. During the early pandemic, women could not have a companion in the postpartum ward. Also, childbirth preparatory classes were closed and midwives were reluctant to offer their services as private midwives. But after one month the situation improved. Virtual preparatory classes became available and private midwives resumed their services supporting women at birth. The fifth wave of the epidemic, caused by the delta variant, resulted in the highest number of infections and deaths in comparison with previous waves in the country. Although a lockdown was imposed, access to health care facilities was not limited and pregnant women could visit hospitals. They were also allowed to have a private midwife in labor and a companion during postpartum. At this juncture in the COVID-19 pandemic in Iran, vaccination of pregnant women had not yet been begun but almost all midwives had been infected and all had been vaccinated.

We explored the role of variables related to labor and birth and also psychological variables in birth satisfaction. Our results indicate that emergency cesarean, instrumental birth, episiotomy, Entonox analgesia, low well-being score, high fear of COVID -19, low satisfaction with pregnancy and a low satisfaction with spouse’s support are predictors of lower levels of birth satisfaction. The overall proportion of the variance in birth satisfaction explained by all variables is 17.4%. Labor and birth variables explained 12.2% of the variance in birth satisfaction.

Our results are in line with those of Preis and colleagues (29). They found that in the pandemic period, established predictors of low birth satisfaction such as nulliparity, mode of birth, social support, and labor and birth complications explained 35% of the variance in birth satisfaction. According to the same study, pandemic-related variables including maternal concerns about preparation for birth and restrictions on the number of family members allowed to accompany a birthing mother explained 3% of the variance in birth satisfaction (29).

We found that fear of contracting COVID-19 during the fifth wave of the disease was a predictor of lower birth satisfaction. But we expected it to be a stronger predictor of lower birth satisfaction than our results suggest. Fear of COVID-19 may induce higher anxiety before admission to the hospital because of uncertainty and unfamiliarity with the hospital setting and the higher risk of contracting COVID. But during labor and birth, uncertainty and unfamiliarity are diminished and women feel quite relieved when they are about to be discharged from the hospital in a few hours. Previous studies have found that the level of stress experienced during pregnancy and childbirth was significantly associated with birth dissatisfaction (15, 18). We found no difference in the levels of fear of COVID-19 between parity or income groups. This implies that all mentioned groups of women experienced fear of COVID-19. In contrast, the results of a previous study indicate that fear of COVID-19 was associated with parity and stage of pregnancy (30).

We found that compared to elective cesarean, emergency cesarean and instrumental birth were better predictors of lower levels of birth satisfaction. This result may be explained by delays and shortcomings in the implementation of policies and guidelines promoting physiological vaginal birth in our setting. Results from previous studies indicated that cesarean was associated with lower birth satisfaction in comparison with physiological vaginal birth (4, 8, 18, 29, 31).

In our study, women’s low level of well-being was a predictor of lower birth satisfaction. We found similar result in our previous study (8). Comparisons of well-being according to parity and income indicate that in the pandemic period, well-being mean scores are not different between parity and income groups. This implies that the pandemic has influenced well-being in both these groups. In the present study, having a pregnancy that is stressful and characterized by hassle was a psychological predictor of birth satisfaction. Also, low satisfaction with spouse’s support could predict birth satisfaction. These results are in agreement with the findings of previous studies (8, 32).

We found that vaginal birth by episiotomy rather than spontaneous tear is a predictor of birth satisfaction. In our sample, 24.8 and 32.1 percent of the women experienced episiotomy and spontaneous tear, respectively. Our study was conducted in a mother friendly hospital and so episiotomy is not performed as a routine procedure; however, because it is a training hospital, newly assigned obstetrics residents may perform episiotomy. In a study in Tehran, Iran, vaginal birth by episiotomy accompanied by tear was a predictor of birth satisfaction (33).

Our results indicate that receiving Entonox analgesia is a predictor of lower birth satisfaction. Entonox analgesia is used for vaginal birth. Our result is not in accord with those reported by Fumagalli and colleagues. Their findings indicate that none of the intrapartum interventions was associated with birth satisfaction (34).

In our study, having a stressful, hassled pregnancy was a predictor of lower birth satisfaction. Also low satisfaction with spouse’s support could predict lower birth satisfaction. These results are in agreement with findings of our previous study (8). In a study on 225 postpartum women in Khaf, Iran, childbirth experience improved with lower hassle and an increased sense of uplift (32).

We found that giving birth to an infant with high birth weight is a predictor of lower birth satisfaction. High birth weight may cause long labor and higher pain levels which are associated with lower levels of birth satisfaction.

We found that none of the socio-demographic variables were related to maternal birth satisfaction. This is in line with the study by Fumagalli et al. (34) and our previous study (8). Results from the present study indicate that primiparity is an obstetrical predictor of lower birth satisfaction. This is in agreement with the results of several previous studies (18, 33, 34). Satisfaction with childbirth is related to three factors: women’s attributes, quality of care received, and stress. Several studies have shown that multiparas experience lower levels of stress and fear of childbirth because of their previous birth experience. Also, giving birth to the first child is usually more difficult than the second or third child. That the satisfaction with current birth is higher in multiparas than primiparas may be the result of their previous experience which decreases their fear of birth and also makes delivery easier and more comfortable.

We found no relationship between the presence of a private midwife during labor and the birth satisfaction score. It seems that the role of private midwives in improving birth satisfaction is not as significant as commonly believed and needs further evaluation. Childbirth preparatory classes have become increasingly popular among Iranian pregnant women. In prenatal visits, pregnant women receive the option to register for free childbirth preparatory classes. During the pandemic, virtual classes were held by midwives who work in private practices or public health centers. After completing the course, participants may opt to have a private midwife during labor, birth, and early postpartum. Private midwives are permitted to support women during active labor, birth, and early postpartum. Most of them do not have permission to perform vaginal exams, delivery of the baby, and other interventions. Obstetrics residents or midwifery students under the supervision of mentors usually perform the deliveries.

We found that women who had a private midwife during labor and birth reported a higher level of fear of COVID-19. A possible reason for this may be that during the pandemic, women with higher levels of fear had hired private midwives to receive better care. But because this is a cross-sectional study, we cannot conclude a causal relationship between the two factors. In a study in Russia, the rate of having a support person at labor decreased from 58% in the pre-pandemic period to 27% in the pandemic due to the COVID-19 restrictions (35). Results of a study in the US indicate that the number of support persons during birth predicted birth satisfaction (29).

### Limitations and strengths

The main limitation of this study is due to its cross-sectional design which makes it difficult to establish cause and effect relationships between some variables. The strong points of this study are the large sample size. We investigated birth satisfaction in the early hours after birth; so, in comparison with studies that were conducted several months after birth, our results are relatively more precise. This study was performed using a sample of postpartum women who gave birth in a conventional birth setting. Its findings cannot be generalized to populations who give birth in labor-delivery-recovery settings where women are isolated from other parturient and therefore are less worried about contacting COVID-19.

### Implications for future research

Data collection for this study was completed before the vaccination of pregnant women against COVID-19 had started. We recommend that further studies be undertaken in Iran to explore the effects of vaccination on birth satisfaction. According to our findings, hiring a private midwife was not associated with higher birth satisfaction. We believe that this finding merits further exploration and so we recommend that qualitative studies be conducted to explore perspectives of women and midwives on the role of private midwives.

## Conclusions

We found that fear of COVID -19 is a predictor of lower birth satisfaction. Our findings also indicate that variables related to labor and birth were predictors of birth satisfaction. These variables which were responsible for a large part of the birth satisfaction during the pandemic include the following: emergency cesarean, instrumental birth, Entonox analgesia, episiotomy, low level of well-being, low satisfaction with pregnancy, and low satisfaction with husband’s support. Based on our findings we recommend a number interventions to increase birth satisfaction and thus maternal mental health. Chief among these are interventions to lower fear of contracting COVID -19 and to reduce rates of episiotomy, emergency cesarean, and instrumental birth.

## Data Availability

All relevant data are within the manuscript and its Supporting Information files.

## Acknowledgements

We would like to thank pregnant women who participated in this study.

